# AIRWAY-XR: Augmented Instruction to Refine Wayfinding and Yielding Skills in Emergency Medicine Residents for Intubation using Mixed Reality Technology

**DOI:** 10.1101/2025.01.06.24319788

**Authors:** Neil Bhavsar, Sandhya Sriram, Shriman Balasubramanian, Christian Davidson, Wojciech Piechowski, Jordan Zimm, Sowmya Sanapala, Alexander Fortenko, Maria Lame, Peter Wagoner Greenwald, Jonathan St. George, Alexandros Sigaras

## Abstract

The goal of this study was to assess if mixed reality technology (MR) is a feasible training tool for educating new learners in endotracheal intubation. This is a feasibility trial to establish the feasibility of an MR airway education module compared to traditional airway teaching. The study participants were twenty-one postgraduate year one (PGY1) physicians accepted to an emergency medicine residency program located in a large, urban setting. The residency program is located in New York City, and has academic affiliations with two large, urban, academic emergency departments (ED) that each treat over seventy thousand patients per year. We enrolled 20 Emergency Medicine (EM) first-year residents into two research arms. Group A consisted of 10 first-year residents who utilized a novel MR education module containing self-guided training and real-time feedback via the Microsoft HoloLens 2. Group B consisted of 10 first year residents who trained on a phone-based module containing the same self-guided content and received real-time training via iPad, which is the traditional didactic format. Both groups had remote Senior EM Residents (PGY3) coaches who provided feedback and direction to the participants. Groups were subsequently assessed by Attending EM Physicians on a 10-point scale and given a post-survey to provide feedback on their experience. There was no difference in scores between the HoloLens 2 users and iPad users (HoloLens score [8.6] v. iPad score [8.5]; p = 0.56) or in completion time between the two groups (HoloLens = [3.4 +/− 0.9 min] v. iPad = [3.3 +/− 1.4 min]; p = 0.45). Out of the Group A participants, 70% rated their overall experience between good and excellent. We concluded that using the HoloLens 2 as a didactic model for intubation is feasible, with possible evidence for noninferiority to traditional didactic models if explored further in a larger standardized trial.

## Introduction

### Intubation Skill in Emergency Medicine

Endotracheal intubation is an important, lifesaving procedure that requires mastery in multiple cognitive domains and is often performed under emergent conditions.^1^ Traditional methods of teaching intubation are resource intensive, requiring the time, materials, physical space, and personnel to provide adequate training.^2^ The increasing complexity and accelerating change in our healthcare system only add to the challenges of effective knowledge translation in medicine, and disruptive events like the recent global pandemic highlight the vulnerability of traditional teaching methods. There is an urgent need to develop more robust and resilient forms of learning that increase didactic opportunities by meeting learners where they live, integrating them seamlessly into daily clinical workflows, and erasing the limitations of physical location by merging digital and physical space to actively address the limitations of current teaching practices. Mixed reality (MR) technology can enhance the learning experience by creating an immersive, interactive, and engaging training environment while preserving the relationship between an instructor and learner.^3^ There have been multiple studies that have evaluated the importance of training emergency physicians to master emergency airways, but very few evaluate the methods of training.^4–5^

### Mixed Reality

Mixed reality (MR) is a technology that merges virtual content into the physical environment, enabling interactive experiences where digital and real-world elements coexist and interact in real time. From a broader perspective, MR has been studied as an educational tool for learners in various industries. When applied in education, MR, provides better learning engagement, more interaction between students and learners, and increased motivation among learners using MR. Additionally, MR can appeal to learners with low spatial abilities who need augmented learning visualization and engaging visual aids to support learning.^6^ MR can create an immersive experience, creating an environment to nurture experiential learning.^5^

### HoloLens 2

An example of an MR capable tool is the Microsoft HoloLens 2 headset.^7^ The Microsoft HoloLens 2 (Figure 1) is an augmented reality headset that combines digital holographic content with the user’s real-world environment. It is a HIPAA compliant MR device capable of audio and video communication in a three-dimensional (3D) setting. The device creates an immersive mixed reality space where users can interact with virtual objects and information through natural gestures such as eye and hand movements. Its spatial mapping capabilities enable precise placement and interaction of persistent holographic elements within the physical surroundings, which are fully visible without disruption. The head-mounted display (HMD) provides the user with an expanded field of view (FOV), enhancing user engagement. The device measures the users interpupillary distance to design a tailored and comfortable experience. The headset is designed for comfort with an adjustable headband ensuring extended wear.

**Figure 1.**
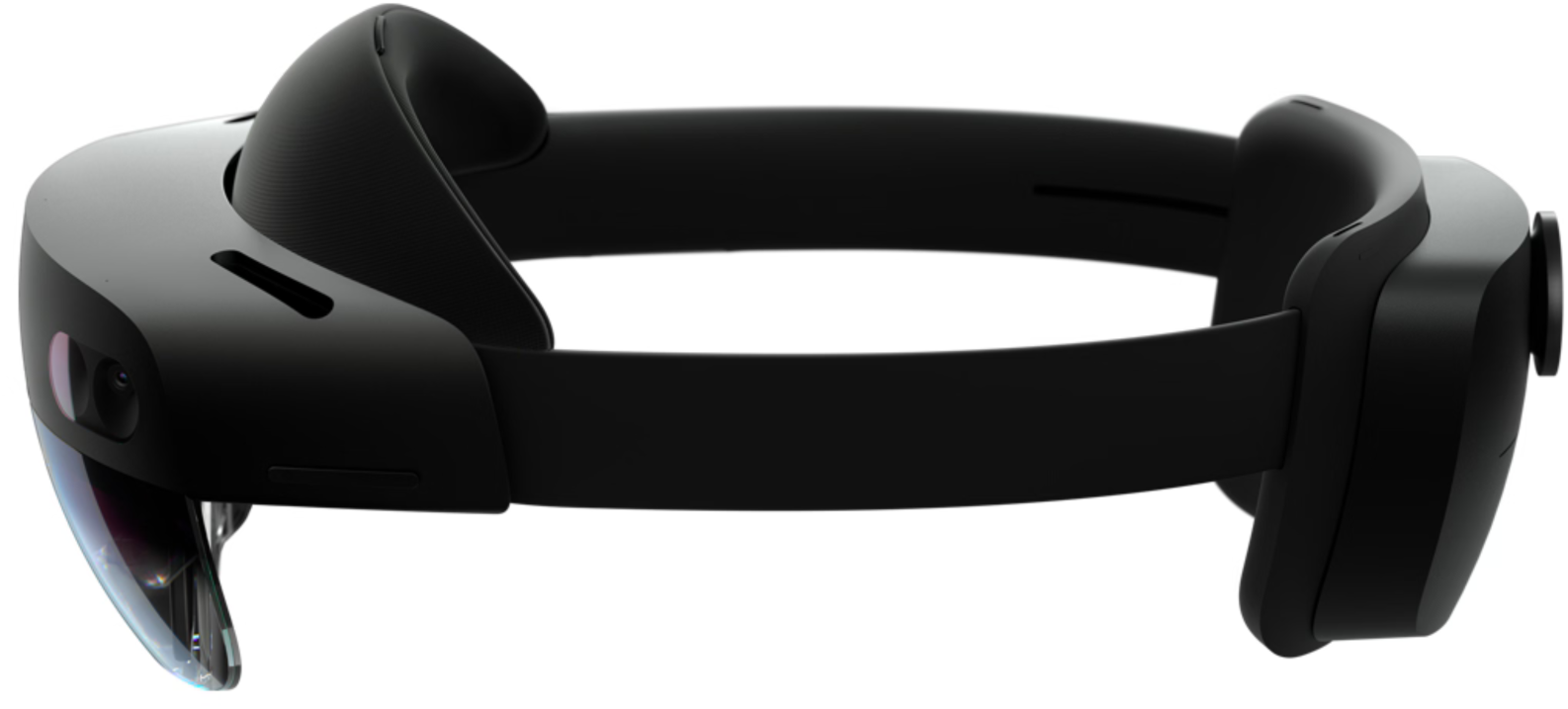
Microsoft HoloLens 2. (HoloLens 2—Overview, Features, and Specs | Microsoft HoloLens [Internet]. www.microsoft.com. Available from: https://www.microsoft.com/en-us/hololens/hardware#document-experiences)

### Microsoft Dynamics 365 Guides

To display the self-guided education module on intubation within the HoloLens 2, we utilized the Microsoft Dynamics 365 Guides application (Guides). This application allows users to both author a step-by-step guide and use it to follow a protocol. Each step involves instructions written on a mixed reality panel, with options to add holographic 3D objects such as arrows or box outlines to display visual information that interacts with real-world objects. Guides instructions can also include a video or image for further clarification on steps. To navigate through the steps, users can use natural hand gestures to select front and back arrows, focus their gaze on the arrows, tap holographic arrows, or use voice commands. Users can also connect to a Microsoft Teams call within the Guides application and receive additional guidance from a remote coach. Guides brings 2D content into an immersive 3D space to improve engagement and learning outcomes.

### The Use of Mixed Reality in Clinical Applications

The HoloLens 2 has already found use in the healthcare setting, such as in surgical navigation in the operating room.^8^ MR provides computer-generated information superimposed to a real-world environment which has resulted in improved accuracy, safety, and efficacy in surgical procedures when trialed by a select group of surgeons.^8^ MR technology has also been used in remote bedside modules, tele-mentoring, and teleconsulting.^9^ Learners can engage with material while coaches and teachers are remote, educating learners with a laptop in a different room.^9^

### Mixed Reality in Medical Education

Many studies show positive feedback that medical education, specifically airway education, is a clear use case for mixed reality. For example, Alismail et al. demonstrated the use of hand-held augmented reality (AR) devices to improve intubation outcomes when scored on a checklist— while participants took a longer time to intubate when using the AR device, they made no mistakes on the checklist, whereas participants who did not intubate with the AR device tended to miss some of the steps.^10^ However, most of the studies that exist currently focus on the actual use of AR or MR, and do not have a real-time coach in a remote location.^9^ The purpose of this study is not only to use the HoloLens 2 for self-guided training, but to give real-time feedback to learners and to allow for real-time education and adjustment to be made. Not only does this allow for learners to build the habits of essential airway management techniques early, but this preserves the important relationship between a coach and a learner, even in a remote setting.

## Methods

This study was implemented and adapted as an educational module established in the Protected Airway Collaborative (PAC) for MR with Group A while making no changes for Group B from standard didactic training in airway education. PAC was designed as a tool to educate novice and experienced physicians in the skills of managing an airway in various scenarios ranging from beginner to advanced skills. The model utilizes short, sixty-second educational videos that are accessible through engaging posters with QR codes. It deploys impactful graphics, embedded multimedia content, and a flipped classroom approach.

The study design is a feasibility experimental study to determine if the Microsoft HoloLens 2 is noninferior to traditional teaching with a remote coach. This study was approved by the Weill Cornell Medical College Institutional Review Board (IRB#23-01025554-02). The focus of the intervention is twofold: (1) demonstrating the use of the Microsoft HoloLens 2 device to watch PAC videos and (2) utilizing MR features to augment the user’s field of view (FOV) including videos, highlights, and remote coaching directly in the FOV.

The study participants consisted of twenty-one post-graduate year one (PGY1) physicians from an Emergency Medicine residency program located in a large, urban setting. The residency program is located in New York City, and has academic affiliations with two large, urban, academic emergency departments (ED) that each treat over seventy thousand patients per year. The PGY1 physicians had not started working clinically in the ED at the time of the study and were undergoing a two-week orientation. Part of their orientation included advanced airway training. Users were coached to intubate mannequins equipped with anatomically realistic upper airway anatomy.

Figure 2 demonstrates the flow of the study design for both groups A and B. The design involved using the self-guided nature of the PAC videos, combined with a virtual coach readily available on a Microsoft Teams call for real-time feedback. The coaches were controlled to be PGY3 emergency medicine residents. They were chosen because of their role as senior residents, part of which involves teaching and coaching junior residents through procedures. The users were randomly divided into two groups and were not blinded due to the nature of the intervention. Both groups completed a pre-intervention survey about their digital literacy skills as well as any prior experience with augmented reality, virtual reality or mixed reality. The control group received the PAC video instruction on their personal devices, intubation practice, and real-time remote coaching via an iPad. The intervention group was onboarded on HoloLens 2 and shown how to interact on Microsoft Guides. Figure 3 shows one such participant undergoing onboarding, during which they interacted with mixed media content on the HoloLens 2 to gain familiarity with the technology in a simple cup stacking exercise.

**Figure 2.**
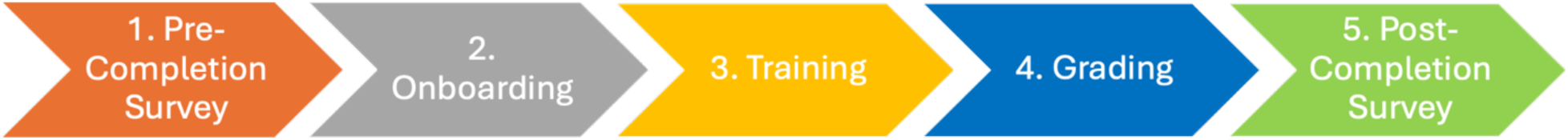
The stages of the Airway MR Study.

**Figure 3.**
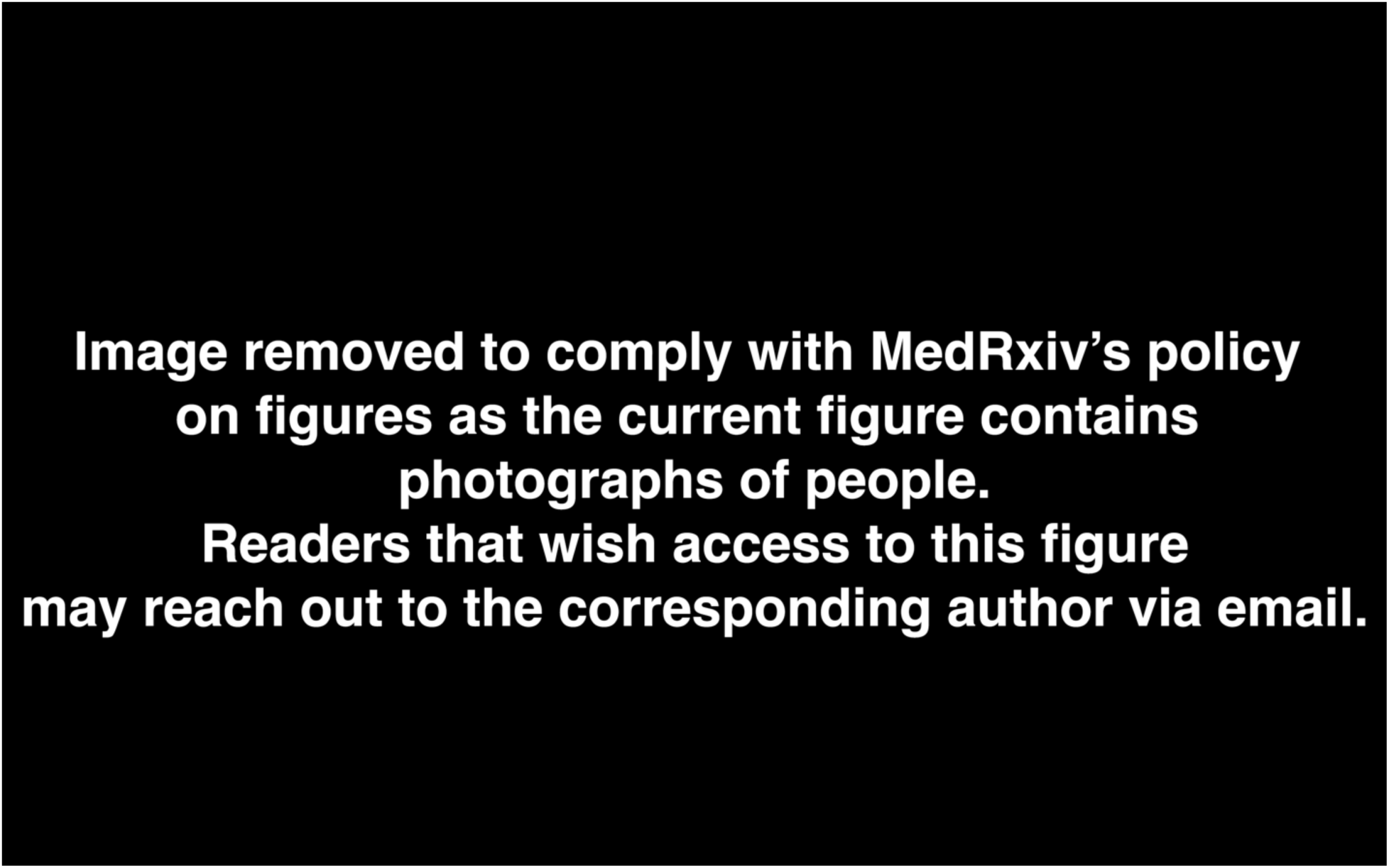
Group A (HoloLens 2) participant undergoing onboarding. The participant is tasked to complete a series of hand eye coordination cognitive tasks by stacking cups into a pyramid.

Group A then received an introduction to the Microsoft HoloLens 2 device and connected to a Microsoft Teams call with the remote coach. Both groups then completed the self-guided PAC video instruction, either through the HoloLens 2 or via a smartphone, as shown in Figure 4. As per the traditional PAC didactic model, Group B completed their self-guided smartphone training without supervision before joining the remote Teams call, unlike Group A who completed the self-guided training while on call with the remote coach.

**Figure 4.**
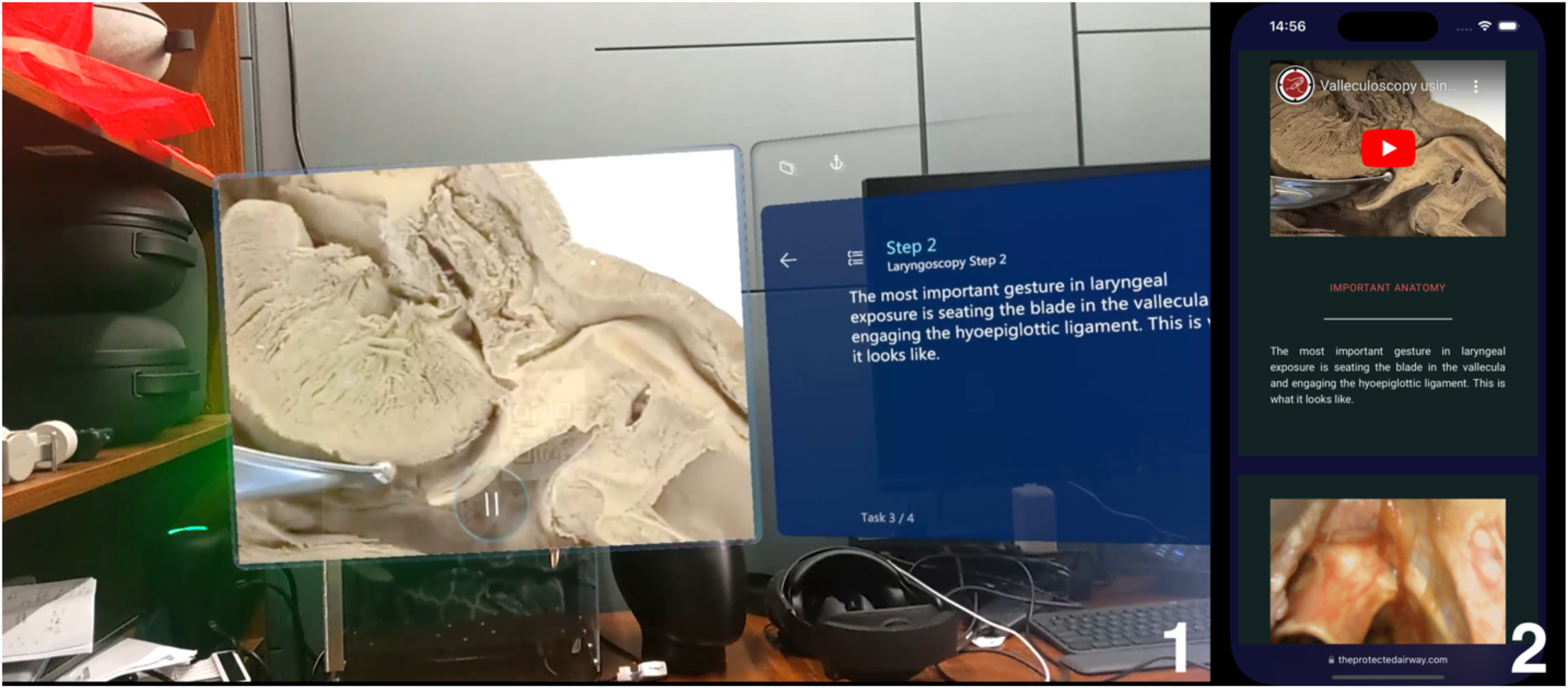
Self-guided PAC video instruction for Group A participants via HoloLens 2 (1) and Group B participants via smartphone (2).

Upon completion of the self-guided training, the remote coaching consisted of real-time instruction from a senior resident, including direct FOV augmentations for the HoloLens 2 group, as shown in Figure 5 (1) and a regular camera video feed on the iPad group (2).

**Figure 5.**
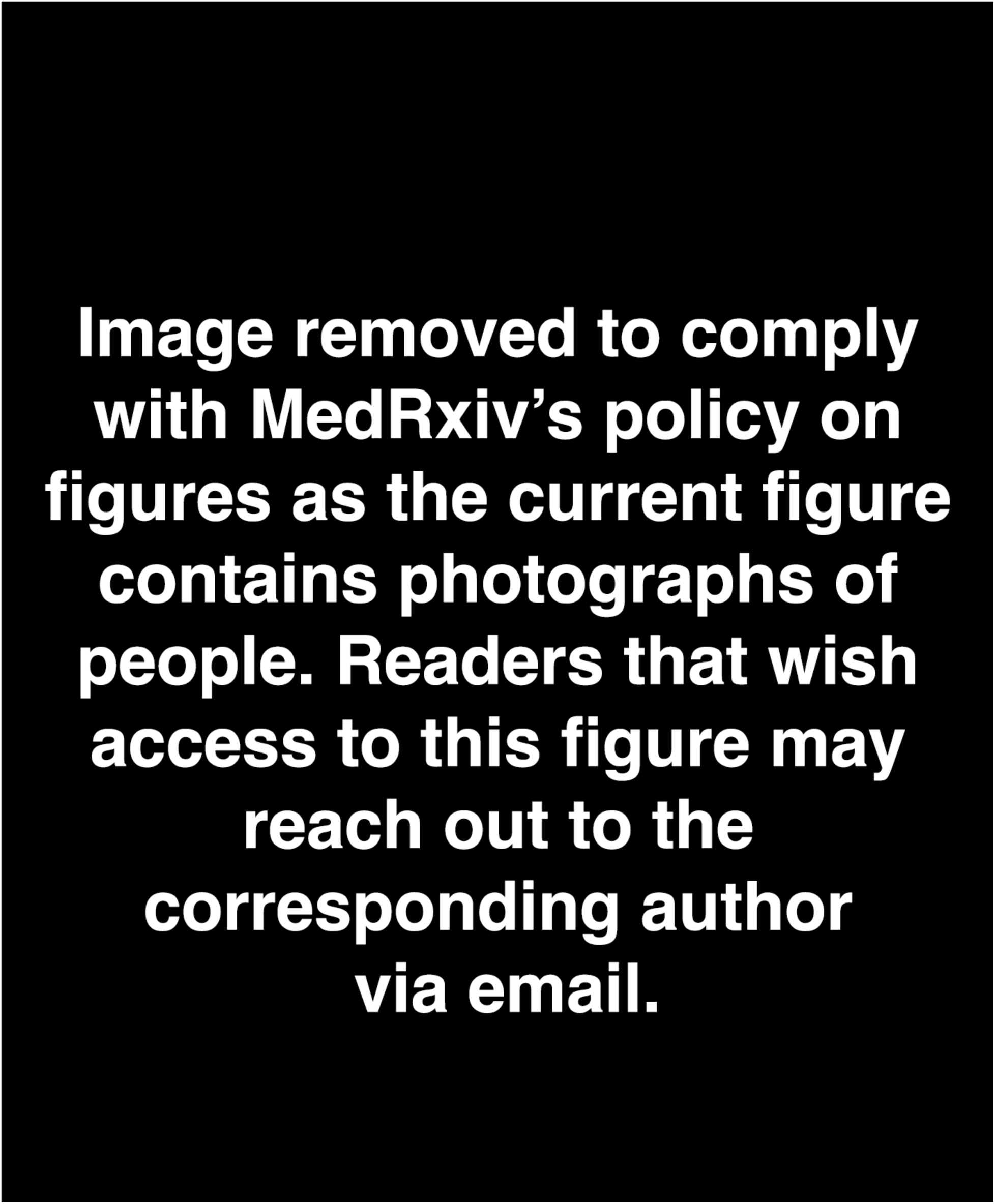
Remote coaching on Group A – HoloLens 2 with direct FOV augementations (1) vs regular camera video feed for Group B – iPad (2).

Next, both groups received assessment of their intubation skills, which was graded on predetermined criteria (Table 1). Each resident attempting intubation was graded by an attending emergency physician. Finally, both groups completed a post-intervention survey to evaluate their onboarding, training, and grading experiences.

**Table 1-.** The grading rubric used to grade all users.

| Action | Points |  |
| --- | --- | --- |
| 1 | Scissoring technique | 1 |
| 2 | Laryngoscope placed midline | 1 |
| 3 | Found uvula | 1 |
| 4 | Blade against base of tongue as pushing forward | 1 |
| 5 | Found Tip of epiglottis | 1 |
| 6 | Advanced blade into vallecula | 1 |
| 7 | Optimize view of vocal cords - (Lifting up appropriately) | 1 |
| 8 | Delivered tube | 1 |
| 9 | Removed Stylet | 1 |
| 10 | Inflated Cuff | 1 |
|  | Points total | 10 |

## Results

The collected data included formal assessments for both groups A and B after the training, as well as time spent while learning. User survey data was also collected to assess the learners’ experiences. There were 10 participants in the control group and 10 participants in the test group. As shown in Figure 6, when Group A was compared to Group B, there was no significant difference between the scores on the assessment (HoloLens 2 score [8.6] v. iPad score [8.5]; p = 0.56). Additionally, there was no significant difference in completion time between the two groups (HoloLens 2 = 3.4 min v. iPad = 3.2min; p = 0.45), as shown in Figure 7. Scores largely ranged from the 8-10 range out of a 10-point system as seen in Figure 8, indicating a strong comprehension of the training and content across both groups.

**Figure 6.**
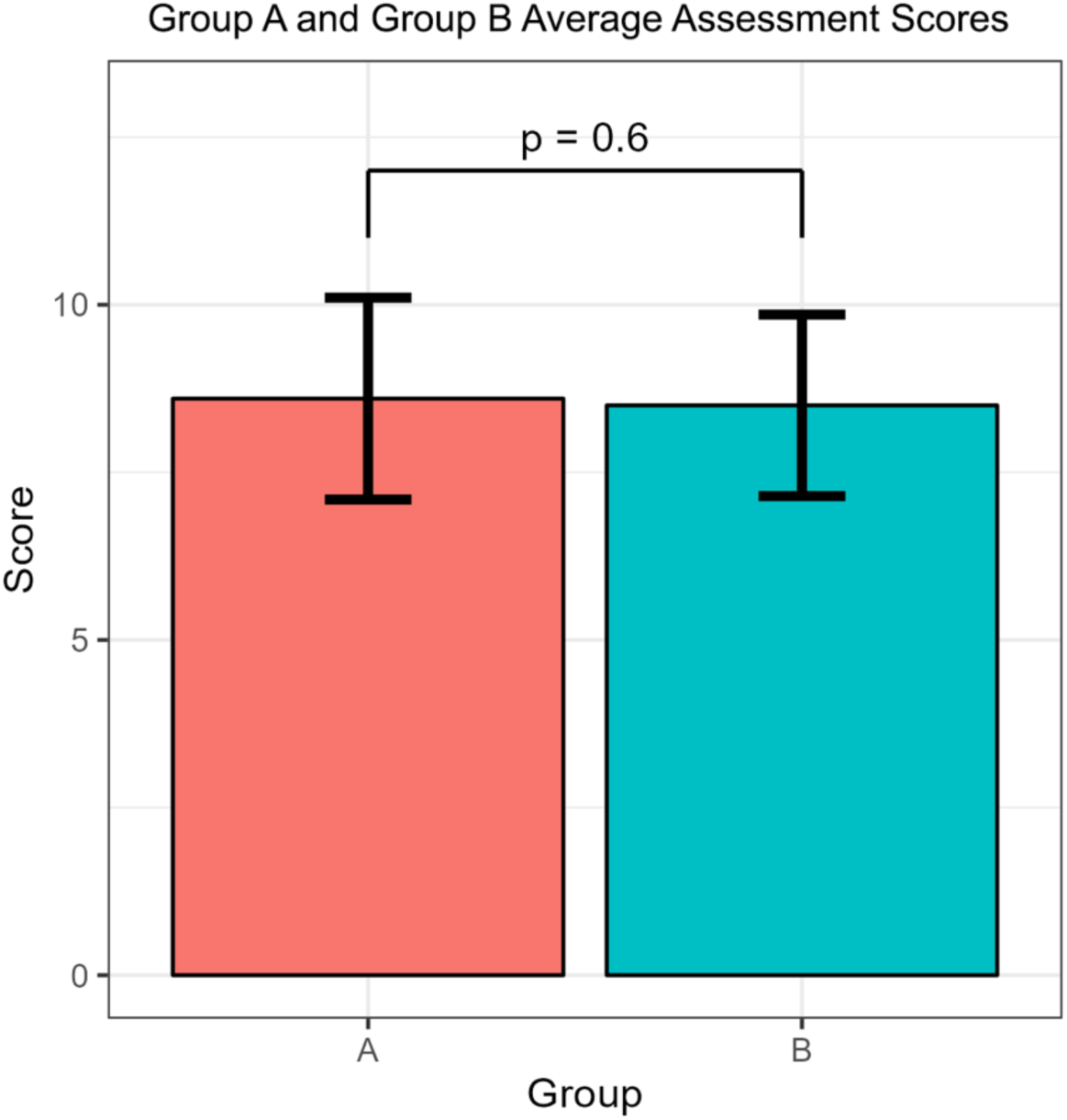
Group A and Group B Average Assessment Scores.

**Figure 7.**
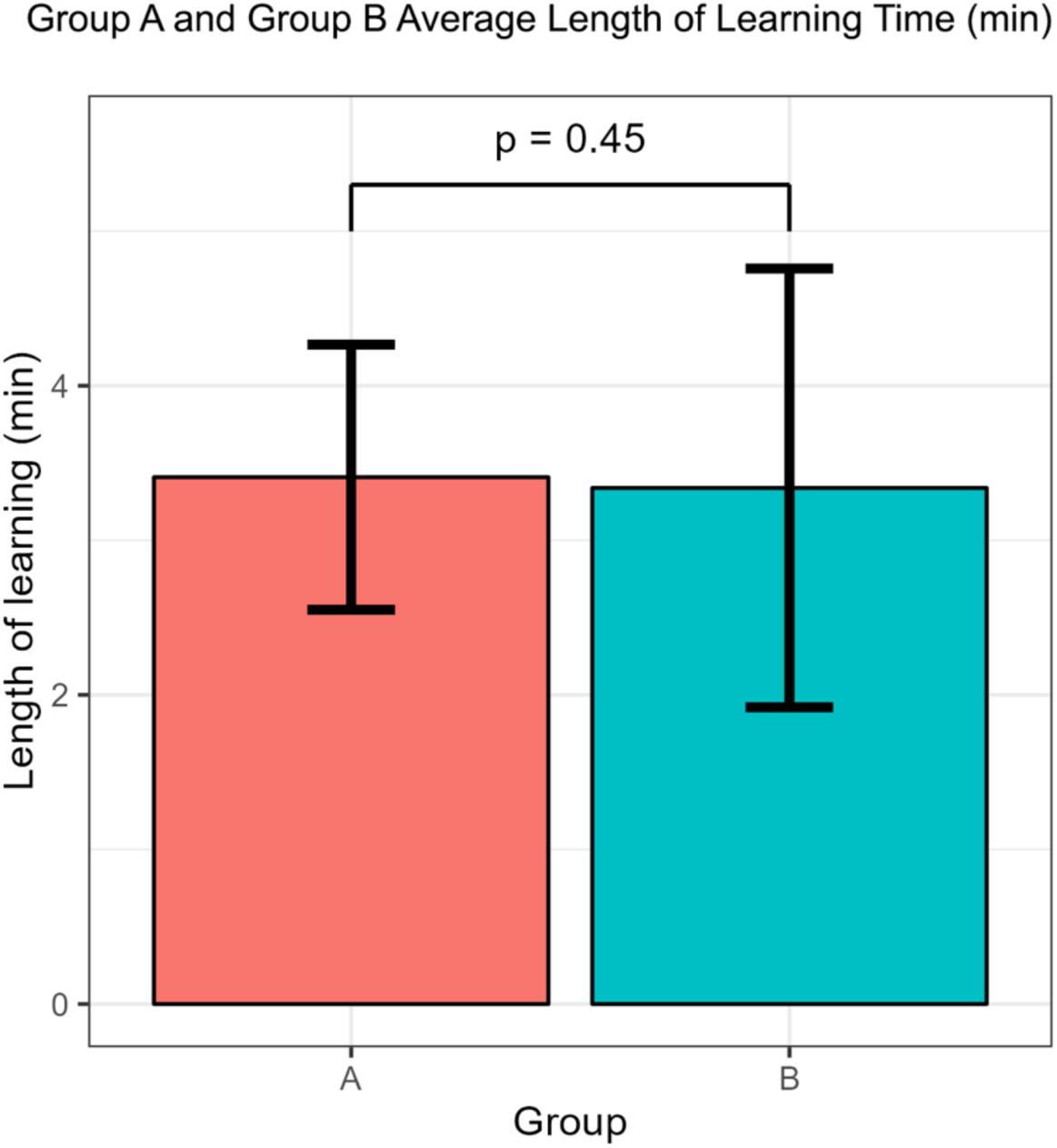
Group A and Group B Average Length of Learning Time (min).

**Figure 8.**
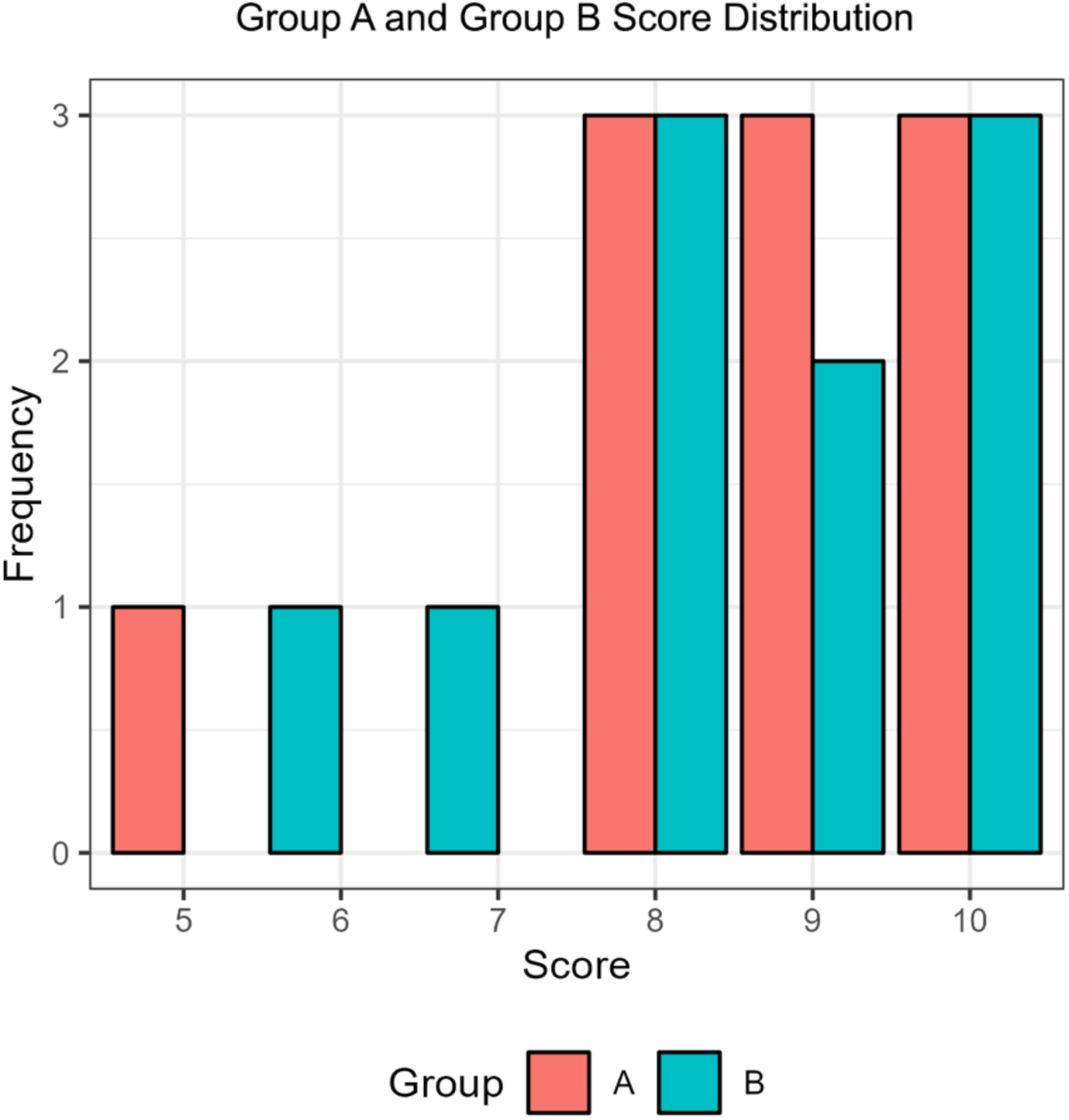
Group A and Group B Score Distribution.

The post-survey results indicate that HoloLens 2 users largely found their experience to be positive, with 70-80% of users giving the highest score when indicating that the remote coaching and HoloLens application was helpful for performing the task, as shown in Figure 9. Experience comparison between Group A and Group B shows that 70% of participants in Group A found the experience to be between Good and Excellent, while all users in Group B found the experience to be between Good and Excellent, as detailed in Figure 10.

**Figure 9.**
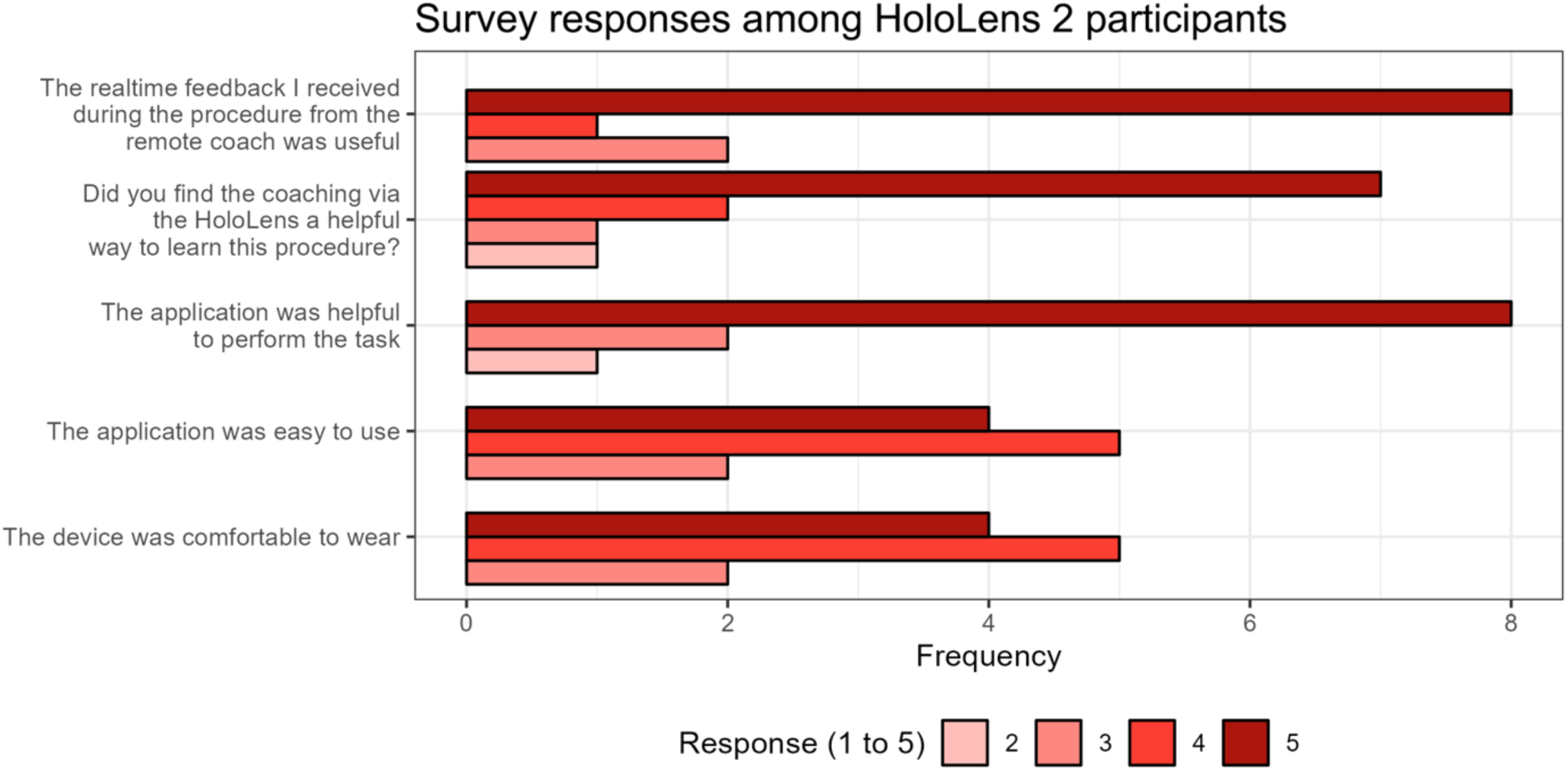
Survey responses among Group A - HoloLens 2 parRcipants.

**Figure 10.**
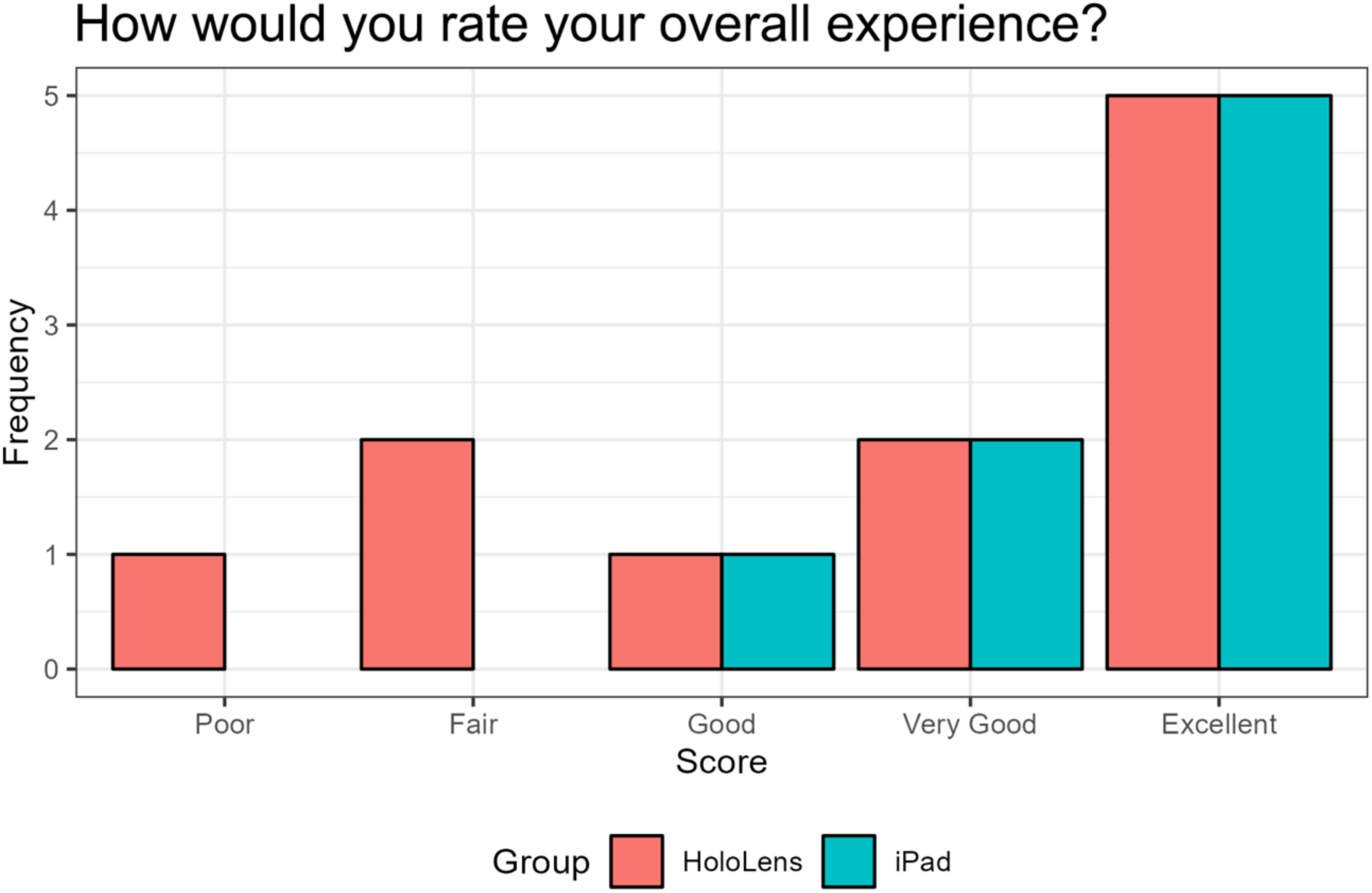
Responses to the question: “How would you rate your overall experience?” from both groups A and B.

## Discussion

### Result Analysis

Our data supports that MR may be a feasible and appropriate modality for teaching and coaching intubation skills to new residents. Group A, the HoloLens 2 group had assessment scores with no difference when compared to traditional videos with a remote iPad coach. The length of learning time between both groups also had no difference. Post-survey data analysis indicates that a majority of users had a positive experience with remote coaching, the comfortability of the headset, and the usability of the application. Three of the ten HoloLens 2 users had a negative experience per the post survey, with two users evaluating the experience as “fair,” while one user evaluated the experience as “poor”. This may be due to technical difficulties with the mixed reality experience, which is explored further in the limitations portion. The role of MR in emergency medicine clinical training will most certainly increase as the technology develops. There is significant potential for remote learning that removes geography as an obstacle. Experts from around the world can coach new learners and provide feedback and visualize the learner’s FOV in real-time.

### Limitations

There were a few limitations to our study. Our sample size was small, as we only had 20 residents enrolled across multiple campuses. There is evidence of feasibility in utilizing HoloLens 2 as an educational tool for intubation; however, given the limitations found during our use, it may be difficult to transition into a clinical environment. There were several instances when the HoloLens 2 would overheat and shut down during the training. The downtime was brief, lasting ten to twenty seconds, and the devices would restart without issue. This problem arose due to overexposure to direct sunlight, which would not be as relevant in a clinical setting, but this would still need to be taken into consideration due to the emergent nature of clinical environments. At times, users also had difficulty hearing the coach due to audio and video communication errors. These errors were also brief, lasting seconds, but pose another challenge when implementing the tool in a clinical environment. Additionally, most first year residents can present with varying levels of experience with intubation. Some of the first-year residents had completed anesthesia rotations, performed emergency intubations in the Emergency Department, or have had careers involving advanced airway experience such as Emergency Medical Services (EMS) or respiratory therapy. On the other hand, some residents have had limited to no experience in intubation. This variation in exposure to the intubation task prior to this study prevented us from assessing noninferiority between the didactic models, and we would need to extend this study to a larger trial with novice participants to truly assess noninferiority or superiority over traditional didactic methods.

### Opportunities for Further Exploration

Currently, we have sufficient evidence to determine feasibility of using MR as an educational tool for intubation, but the study would need to be expanded to encompass a larger number of participants who are all novices in intubation to determine noninferiority against traditional didactics with the iPad. This would prevent varying levels of previous intubation experience from confounding the scoring data and allow us to analyze the effectiveness of the training tool for a broad audience. Survey results from this study showed a positive experience and quick acclimation with the HoloLens 2 onboarding process, so all participants in the experimental group should undergo this same onboarding. To assess skill, increase between pre- and post-training, all participants should attempt an intubation and be scored on the ten-point rubric shown in Table 1 before being provided training. Additionally, all senior residents conducting the remote training portion should be provided with a script to prevent communication-related bias. Each group can then be scored post-training on the same rubric, and both the average pre- and post-training scores and the delta increase can be compared to determine noninferiority or superiority. Metrics on communication levels between participants and coaches in the remote coaching portion can also be evaluated to determine if HoloLens 2 leads to more active conversation and engagement with the coach.

### Implications of Mixed Reality

Mixed reality didactic methods have the potential to improve access to medical education in areas with limited resources by expanding the reach of professional instructors. The technology also comes with significant clinical improvements with each new generation of the device, implying future models may lead to better implementation for clinical education.^11^ Previous studies have supported the use of MR to teach procedural skills to remote learners when compared with traditional didactic techniques, and our results add to the small but growing body of evidence that MR is effective for such purposes.^12–13^ While MR can help democratize access to some educational resources, reliable internet access remains a prerequisite for effective MR teaching. Accordingly, improving global access to the internet will similarly improve the reach of MR to areas that may have fewer professional educators or specialists to teach trainees.

The need for effective remote medical education tools, like MR, was further underscored by social distancing requirements imposed by the COVID-19 pandemic. For a several year period, regulations on travel and trainees severely limited traditional in-person clinical experiences.^14,16–17^ While limiting exposure was crucial in slowing the spread of COVID-19, this period was likely even more detrimental for trainees in resource-poorer areas that may rely on extramural rotations or visiting professors to learn specific skillsets. A functional, widespread MR system could have helped to mitigate some of the educational detriments of the pandemic. Regardless, MR was valuable in addressing several novel challenges that came with COVID-19, including telehealth, drug discovery, patient assessment, mental health management, entertainment, business and industry, and some educational initiatives.^15,16,17^ Many healthcare professionals were asked to perform skills outside their typical repertoire during the COVID-19 pandemic, and MR was shown to effectively promote adherence to a standard intubation checklist in a small group of mixed educational background participants, which included non-physician learners like respiratory therapists and a sleep technologist (Alismail et al.,) our findings of non-inferiority underscore the potential of MR to facilitate skills acquisition.^10^ In cases where geographic distance, isolation from infectious disease, or need for accelerated learning arise, MR has the potential to fill a critical need. It would be positioned to do so more efficiently with early investment and adoption.

The applications of MR for medical training could also be expanded to include non-physician emergency personnel from different training backgrounds. Small groups of emergency medical services (EMS) trainees have already learned via curriculums that include MR, both longitudinally and for specific low frequency events, like pediatric emergencies.^10,17^ Applications have also been developed for disaster management teams to brief first responders using MR before entering a disaster site.^18^ A small study on the use of MR to assist in the “preparedness” and “response” phases of a simulated major disaster response elicited favorable responses from learners; however, such technology has yet to be used to prepare EMS workers in the context a true disaster scenario^18^. Models have similarly been developed to employ MR to assist search and rescue efforts in rubble by reconstructing hypothetical disaster sites with ground-penetrating radar.^19^ Expanding the feasibility of MR even further to mobile devices, one could imagine a remote operator assisting a bystander to perform stabilizing first aid maneuvers superimposed over video streaming via the same phone. ^20^ Developing a way to connect 911 callers with experienced EMS providers--or even emergency physicians--could have a profound impact on the way our EMS system works, especially in time-sensitive situations like cardiac arrest or triage of mass casualty incidents.

## Conclusion

This study demonstrates that mixed reality as an educational tool for developing educational skills in new learners is feasible. All learners enrolled were new residents who were trained in MR several minutes prior to clinical skill acquisition. The results support the use of mixed reality as a tool for more engagement from both the student and teacher. There was no significant difference in the assessment and length of learning for both groups A and B. While some participants in group A experienced disruptions with the HoloLens 2 that impacted their experience, the majority of users felt positively about their experience learning with the device. MR education allows for immersive simulation for learners and real-time FOV visualization for educators, providing point-by-point feedback and instant correction of technique. We can conclude that MR is a feasible method to achieve skill acquisition in intubation for new learners; however, we need to address headset durability, and telecommunication factors, and user comfort for prolonged use before deploying these tools in the emergent clinical setting.

## Data Availability

All data produced in the present work are contained in the manuscript.

## Acknowledgments

The authors would like to recognize the generosity of Caroline Curry and her family in supporting the AI-XR Lab and providing all necessary funding to purchase XR devices to conduct this study. AS would like to thank Olivier Elemento and Andrea Sboner for their invaluable feedback throughout the duration of the project. AS would also like to thank Microsoft and more specifically Erica Capozzi and Brette Bossick for making all the appropriate connections with the Microsoft HoloLens team.

## Author Contributions

Author contributions following CRediT Taxonomy

Conceptualization: NB, SSriram, SB, CD, WP, JM, ML, PWG, AF, JSG, AS

Data curation: NB, SSriram, SB, CD, WP, JM, AF, JSG, AS

Formal analysis: NB, SSriram, SB, CD, WP, JM, ML, PWG, AF, JSG, AS

Funding acquisition: AS

Investigation: NB, SSriram, SB, CD, WP, JM, ML, AF, JSG, AS

Methodology: NB, SSriram, SB, CD, WP, JM, ML, PWG, AF, JSG, AS

Project administration: SSriram, WP, ML, AF, JSG, AS

Resources: ML, PWG, AF, JSG, AS

Software: SSriram, SB, JZ, AS

Supervision: PWG, AF, JSG, AS

Validation: NB, SSriram, SB, CD, WP, JM, SS, ML, PWG, AF, JSG, AS

Visualization: NB, SSriram, JM, AF, JSG, AS

Writing - original draft: NB, SSriram, SB, CD, JM, AF, JSG, AS

Writing - review & editing: NB, SSriram, SB, CD, WP, JM, SS, ML, PWG, AF, JSG, AS

